# iEEG-recon: A Fast and Scalable Pipeline for Accurate Reconstruction of Intracranial Electrodes and Implantable Devices

**DOI:** 10.1101/2023.06.12.23291286

**Authors:** Alfredo Lucas, Brittany H. Scheid, Akash R. Pattnaik, Ryan Gallagher, Marissa Mojena, Ashley Tranquille, Brian Prager, Ezequiel Gleichgerrcht, Ruxue Gong, Brian Litt, Kathryn A. Davis, Sandhitsu Das, Joel M. Stein, Nishant Sinha

## Abstract

**Background:** Collaboration between epilepsy centers is essential to integrate multimodal data for epilepsy research. Scalable tools for rapid and reproducible data analysis facilitate multicenter data integration and harmonization. Clinicians use intracranial EEG (iEEG) in conjunction with non-invasive brain imaging to identify epileptic networks and target therapy for drug-resistant epilepsy cases. Our goal was to promote ongoing and future collaboration by automating the process of “electrode reconstruction,” which involves the labeling, registration, and assignment of iEEG electrode coordinates on neuroimaging. These tasks are still performed manually in many epilepsy centers. We developed a standalone, modular pipeline that performs electrode reconstruction. We demonstrate our tool’s compatibility with clinical and research workflows and its scalability on cloud platforms.

**Methods:** We created *IEEG-recon*, a scalable electrode reconstruction pipeline for semi-automatic iEEG annotation, rapid image registration, and electrode assignment on brain MRIs. Its modular architecture includes three modules: a clinical module for electrode labeling and localization, and a research module for automated data processing and electrode contact assignment. To ensure accessibility for users with limited programming and imaging expertise, we packaged iEEG-recon in a containerized format that allows integration into clinical workflows. We propose a cloud-based implementation of iEEG-recon, and test our pipeline on data from 132 patients at two epilepsy centers using retrospective and prospective cohorts.

**Results:** We used iEEG-recon to accurately reconstruct electrodes in both electrocorticography (ECoG) and stereoelectroencephalography (SEEG) cases with a 10 minute running time per case, and ∼20 min for semi-automatic electrode labeling. iEEG-recon generates quality assurance reports and visualizations to support epilepsy surgery discussions. Reconstruction outputs from the clinical module were radiologically validated through pre- and post-implant T1-MRI visual inspections. Our use of ANTsPyNet deep learning approach for brain segmentation and electrode classification was consistent with the widely used Freesurfer segmentation.

**Discussion:** iEEG-recon is a valuable tool for automating reconstruction of iEEG electrodes and implantable devices on brain MRI, promoting efficient data analysis, and integration into clinical workflows. The tool’s accuracy, speed, and compatibility with cloud platforms make it a useful resource for epilepsy centers worldwide. Comprehensive documentation is available at https://ieeg-recon.readthedocs.io/en/latest/

**Graphical Abstract:** 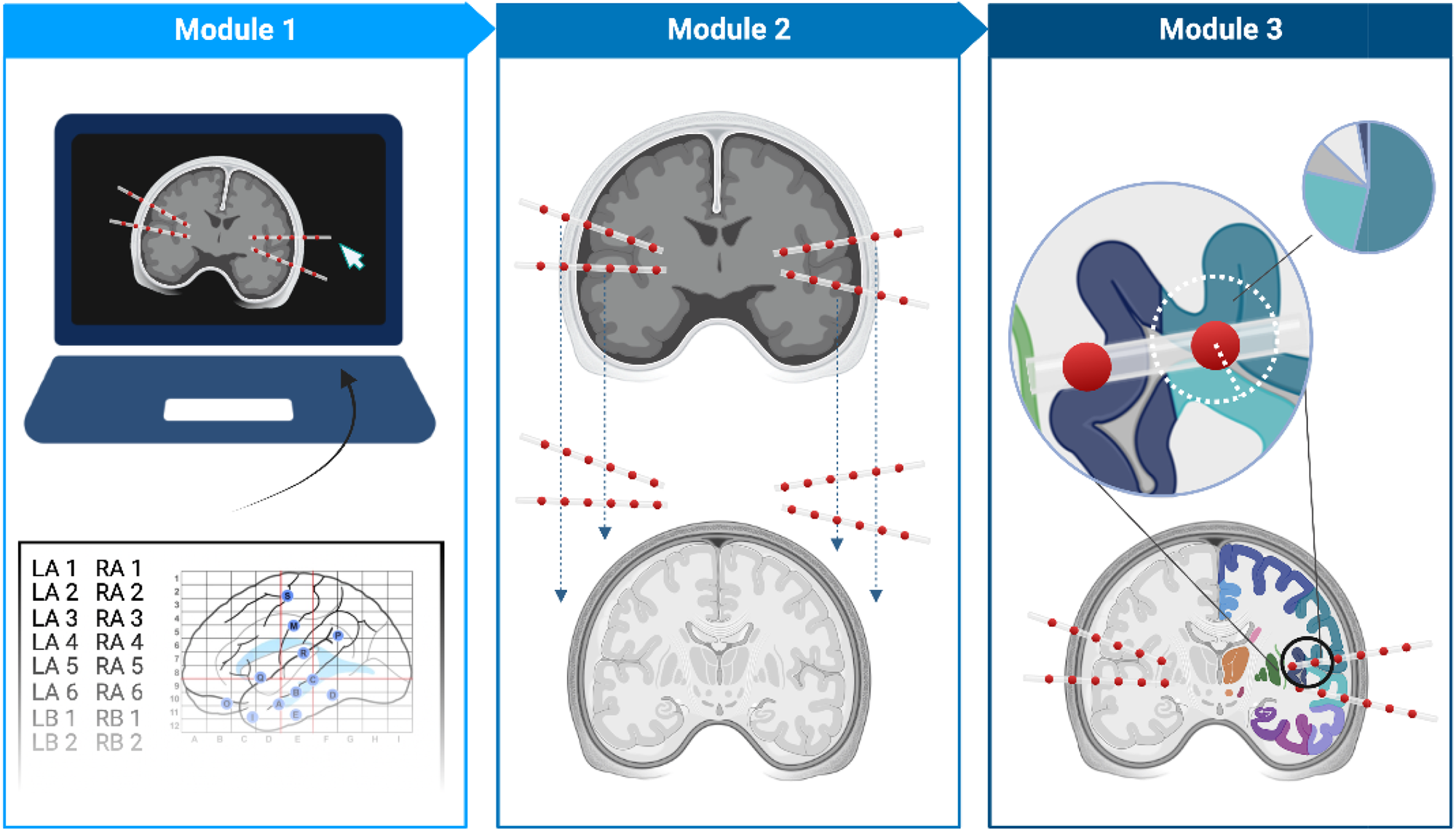

## Introduction

Epilepsy is a neurological disorder that affects around 70 million individuals worldwide, with 30% of patients having drug-resistance and experiencing chronic, uncontrolled seizures^1^. To determine the optimal treatment plan, it is essential to precisely localize epileptic networks that are responsible for seizures. Clinicians apply a combination of non-invasive and invasive techniques, such as magnetic resonance imaging (MRI) and intracranial electroencephalography (iEEG), to localize epileptic networks. MRI can identify abnormal brain regions with high spatial resolution, while iEEG provides a direct measurement of brain activity with high temporal resolution. iEEG involves placing electrodes invasively in the brain. After implantation, confirming the electrodes’ anatomical loci is necessary for several reasons. First, it allows for a comprehensive evaluation of which brain areas will be effectively sampled during the invasive study, which is important to interpret interictal and ictal patterns and to understand variability in signal properties (e.g. gray vs. white matter, contacts outside brain parenchyma, and so forth). Second, since brain swelling may occur as a result of implantation surgery, it can be used to quantify electrode shift by comparing their final location with their intended targets. Third, an accurate anatomical map of electrode localization is critical to plan and interpret electrical stimulation ictal and functional mapping.

To confirm the location of iEEG contacts post-surgically, the most common approach is to align (co-register) an anatomical MRI and a post-implant CT scan. This combination provides excellent balance between accurate localization of electrodes (post-implant CT) with high-resolution anatomy without surgical distortion or hardware artifacts (anatomical MRI). Furthermore, it is often of interest to identify the brain region in which the electrode contacts are located, which can be achieved systematically and reliably through the use of an MRI-based brain atlas. In this work, we use the term “electrode reconstruction” to refer to the workflow of electrode labeling, CT and MRI co-registration, and atlas-based electrode contact assignment to brain regions. There is a critical need to develop rapid, accurate, and scalable iEEG electrode reconstruction tools that clinicians can use in their routine workflow.

Various techniques have been proposed for iEEG electrode reconstruction^2–7^. While these methods have been useful in research for determining brain areas that produce epileptiform activities and assessing overlap with the location of surgery, their clinical adoption remains limited. The slow adoption is primarily due to three challenges. *First*, available tools have a steep learning curve for users unfamiliar with programming and image processing. *Second*, current tools are often time-consuming (∼4+ hours runtime), and require constant user input, which makes them impractical for high-volume centers or large multicenter prospective clinical trials. Scalability necessitates consistent and validated pipelines that can process hundreds of cases on both centralized and federated platforms, while ensuring appropriate data harmonization. *Finally*, the currently available pipelines do not leverage recent deep learning-based improvements in image registration and segmentation techniques, which reduce long runtimes and computational requirements, while increasing accessibility.

In this study, we developed and validated iEEG-recon, a standalone pipeline for iEEG electrode reconstruction. We achieved this by a) semi-automatically marking the electrodes on post-implant computer tomography (CT) images, b) co-registering post-implant CT to pre-implant MRI using state-of-the-art rapid brain segmentation and co-registration techniques, and c) incorporating a modular, scalable design consisting of core modules for clinical needs and research modules for flexible parameter tuning. We developed versions of our pipeline compatible with MATLAB and Python programming environments, as well as a stand-alone containerized tool that can be deployed on cloud-based infrastructure. We validated our pipeline retrospectively on extensive patient datasets containing both stereotactic EEG (SEEG) and electrocorticography (ECoG) from post-implant MRI, then tested its viability on data collected prospectively from two level-4 U.S. epilepsy centers. For each run, iEEG-recon generates a quality assurance report and visualizations that can facilitate discussion in epilepsy surgery meetings. Our pipeline is fast, scalable to process hundreds of patient datasets, reproducible, and available as an open-access tool for wider adoption.

## Methods

### Participants

We included 132 patients with drug-resistant epilepsy from two epilepsy centers: the Hospital University of Pennsylvania (HUP: n = 109) and the Medical University of South Carolina (MUSC: n = 23). Our methods were developed and validated on a retrospective cohort of 118 patients (HUP: n = 98 and MUSC: n = 20) and implemented for prospective testing on 14 patients (HUP: n = 11 and MUSC: n = 3). Patients were enrolled serially between 2015 and 2023 after providing written informed consent for iEEG data analysis, in line with the University of Pennsylvania’s IRB-approved protocol (reference number 821778). All patients underwent whole-brain MRI and iEEG implantation (ECoG: n = 23 or SEEG: n = 75), followed by post-implant CT scans. All patients had a post-implant MRI (n = 98). The *retrospective cohort* had detailed clinical annotations from presurgical evaluation meetings and follow-up records on surgical outcomes. Multiple surgical procedures were represented in our dataset, including ablation (N = 33), surgical resection (N=34), responsive neurostimulation (N=9), deep brain stimulation (N=1), vagus nerve stimulation (N=1), and no surgery to date (N=18). The *prospective cohort* data were processed before epilepsy surgery meetings by clinical coordinators and fellows to assess ease of use in routine clinical workflows. Feedback from an expert neuroradiologist (J.S.) was obtained to improve clinical reporting; however, iEEG_recon outputs were not used in clinical decision-making. Subject demographics and clinical characteristics are summarized in **Table 1**.

**Table 1.** Subject demographics and clinical characteristics.

Subject Demographics
| Characteristic | Number of Subjects |
| --- | --- |
| Total Subjects | 98 |
| Age | 34±11 |
| Female | 42 |
| Disease Duration (years) | 15±13 |
| Lesional MRI | 40 |
| Disease Laterality |  |
| *Left* | 47 |
| *Right* | 32 |
| *Bilateral* | 11 |
| *Unknown* | 8 |
| Type of Implant |  |
| *ECoG* | 23 |
| *SEEG* | 75 |
| Type of Surgery |  |
| *Ablation* | 33 |
| *Resection* | 34 |
| *RNS* | 9 |
| *DBS* | 2 |
| *VNS* | 2 |
| *No Surgery* | 18 |

#### Image Acquisition

At both centers (HUP and MUSC), MRI data were collected on a 3T Siemens Magnetom Trio scanner using a 32-channel phased-array head coil prior to electrode implantation. Anatomical images were acquired using a magnetization prepared rapid gradient echo (MPRAGE) T1-weighted sequence (HUP: repetition time = 1810ms, echo time = 3.51ms, field of view = 240mm, resolution = 0.94x0.94x1.0 mm3; MUSC: repetition time = 1900ms, echo time = 2.36ms, field of view = 256mm, resolution = 1.0x1.0x1.0 mm3). Following electrode implantation, spiral CT images (Siemens) were obtained clinically for the purposes of electrode localization. Both bone and tissue windows were obtained (120kV, 300mA, axial slice thickness = 1.0mm, same for both institutions).

#### Image Processing

An overview of the implant reconstruction pipeline is shown in **Figure 1**. We separated our tool into three sequential modules to allow users to choose the level of processing for their use case.

**Figure 1.**
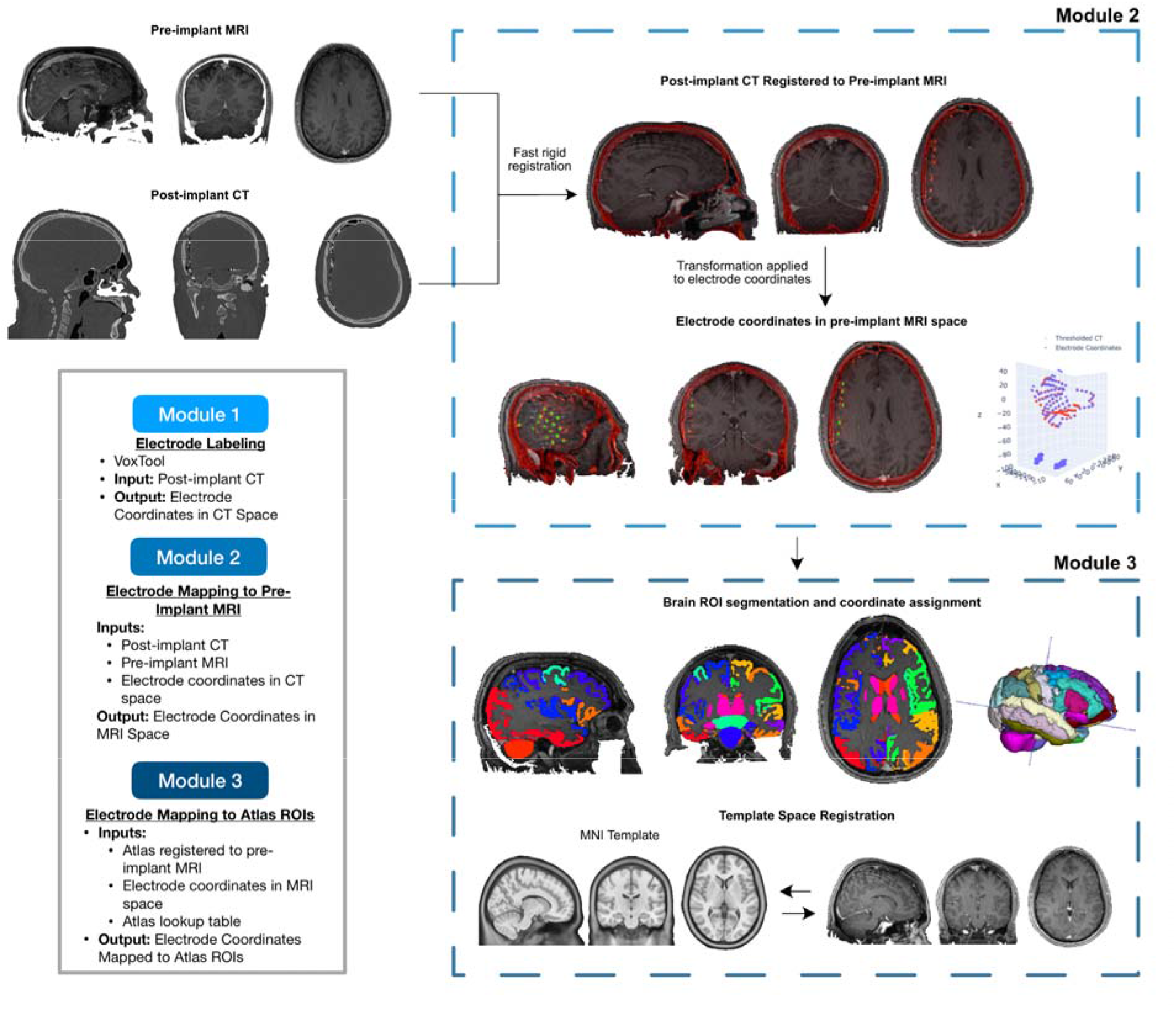
The iEEG-recon pipeline offers a comprehensive solution for electrode marking and reconstruction. By leveraging semi-automatic electrode identification on post-implant computer tomography (CT) images (Module 1), it simplifies and accelerates the process. The pipeline further enhances accuracy by co-registering the post-implant CT with pre-implant MRI using state-of-the-art rapid brain segmentation and co-registration techniques. Built with versatility in mind, iEEG-recon encompasses both core modules tailored for clinical needs (Module 2) and research modules (Module 3) for flexible parameter tuning. This stand-alone and containerized tool can be effortlessly deployed on cloud-based infrastructure. Such adaptability facilitates its integration into multi-center prospective clinical trials, expanding its potential impact.

### Module 1 – VoxTool for Electrode Labeling

In our pipeline, electrode labeling is conducted using VoxTool, a user-friendly graphical user interface for electrode labeling. This software was developed in collaboration with the Penn Memory Lab (https://memory.psych.upenn.edu/Main_Page). Briefly, post-implant CT scans are loaded into VoxTool, which applies an intensity threshold to accentuate the electrodes. The user manually enters electrode labels, then navigates a 3D viewer where electrode voxels can be selected by directly clicking on them. VoxTool is a semi-automatic tool designed to streamline the electrode labeling process. By interpolating electrode labels for grid, strip, and depth electrodes, it accelerates the task while maintaining precision. We have created a video tutorial that demonstrates how to use VoxTool for loading the 3D graphical user interface (GUI) and semi-automatically annotating electrodes. To access the tutorial, please follow this link: https://github.com/penn-cnt/ieeg-recon/tree/main/voxTool.

### Module 2 – Post-implant CT to Pre-Implant MRI Registration

The objective of Module 2 is to register the post-implant CT to the pre-implant MRI. To ensure appropriate skull matching between the CT and MRI acquisitions, we first threshold the intensity of the post-implant CT scan such that only the skull and electrode contacts are visible. After CT thresholding, a rigid registration procedure with 6 degrees of freedom (3 rotation and 3 translation) is applied between the thresholded post-implant CT and the pre-implant MRI. Since brain and skull shapes should not change drastically pre- and post-implantation, 6 degrees of freedom are sufficient for accurate registration^3, 4, 6^. Our pipeline has the option to use either FLIRT^8, 9^ or Greedy^10^ (https://github.com/pyushkevich/greedy) for registration. FLIRT parameters are: 640 histogram bins to discretize the intensity values of the images being registered, and a mutual information cost function with 6 degrees of freedom for registration. For Greedy, we initialize the registration at the center of mass of each image and apply a multi-resolution registration process with 100 iterations at the highest resolution and 100 iterations at the second highest resolution. We stopped the registration after the second highest resolution stage as we did not see significant improvements in registration accuracy by including additional lower resolution levels, but runtime increased drastically. Greedy registration utilizes normalized mutual information as the cost-function with 6 degrees of freedom. We recommend using Greedy as it is faster, but in cases where it fails, FLIRT is available as a fallback option.

The post-implant CT to pre-implant MRI registration generates a transformation matrix which transforms the original electrode coordinates from the post-implant CT space to the pre-implant MRI space. The results of the registration are compiled in an HTML report for quality assurance. For visualization of reconstructed electrodes, an interactive workspace file is automatically generated for visualization in ITK-Snap^11^ (**Figure 2**). ITK-Snap (www.itksnap.org) is an open-source application to visualize and manipulate biomedical images. These visualizations are necessary for manual quality assurance, and any residual rotation between scans could be easily corrected within this interface.

**Figure 2.**
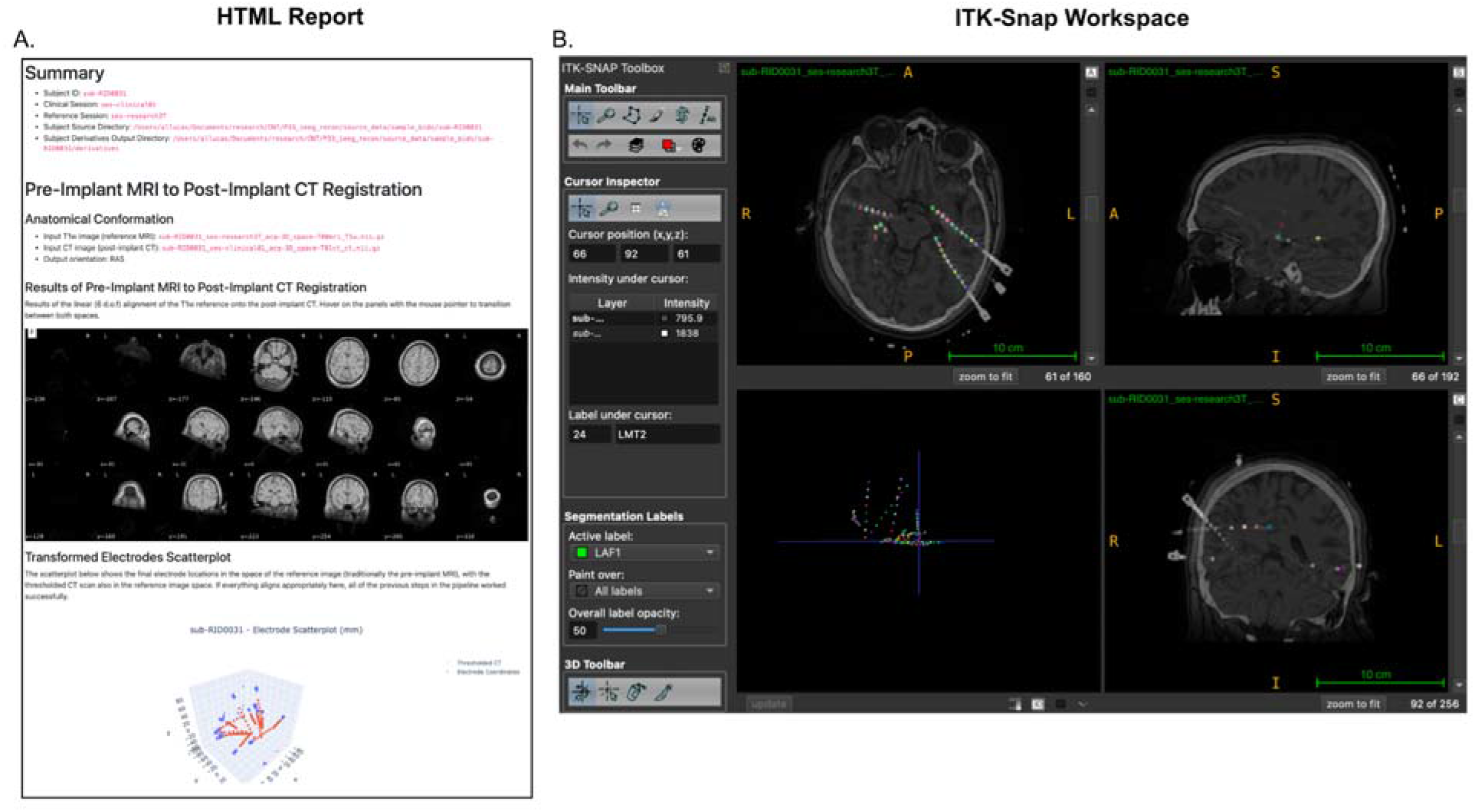
Quality Control Files for Module 2: The output of Module 2 generates an HTML report (A.) that shows the accuracy of the registration between the pre-implant MRI and the post-implant CT, as well as a 3-dimensional scatterplot of the electrodes from the thresholded CT scan and their manually labeled coordinates in the pre-implant MRI space. *ieeg_recon* also generates an ITK-Snap workspace file that overlays the thresholded post-implant CT and pre-implant MRI in the pre-implant MRI space, and plots the coordinates of each of the electrodes as well as their electrode labels provided by VoxTool. This workspace allows for interactive visualization and quality assurance of the electrodes and their locations.

While this process is described for registering the post-implant CT to the pre-implant MRI as a reference, the same process can be applied to any other pair of images (MRI or CT) if one is chosen as a reference. For example, our pipeline also works if the post-implant MRI is registered to the pre-implant MRI, or the post-surgical MRI is registered to the pre-implant MRI (see below).

### Module 3 – Electrode Region-of-Interest Assignment

#### Pre-implant MRI Segmentation

The brain can be subdivided into contiguous regions of interest (ROIs) based on the structural or functional similarity within each region. Assigning electrodes to specific ROIs is useful for confirming iEEG targets^12^. To assign electrodes to a specific ROI, we must identify ROIs on the reference pre-implant MRI. Our pipeline can take as an input any atlas–a particular parcellation scheme–that is in the same space as the reference pre-implant MRI, including: (1) cortical parcellations and subcortical segmentations from either the Desikan-Killany-Tourville (DKT) atlas^13, 14^ or the Taliarach atlas^15^ from FreeSurfer; (2) standard atlases in MNI space (AAL^16^, Schaffer^17^, etc) pre-registered to the reference pre-implant MRI; (3) outputs from more specialized subcortical segmentation approaches such as ASHS^18^ and THOMAS^19^. Our pipeline enables users to choose any atlas for electrode assignments and encourages application of multiple atlases for robustness^20^.

#### ANTsPyNet Segmentation

IEEG-recon has an option to generate and use a cortical, subcortical DKT parcellation, and tissue segmentation (DeepAtropos), from pre-implant MRI using ANTsPyNet^21^. We included ANTsPyNet, as opposed to FreeSurfer, due to its speed that allows obtaining both cortical and subcortical segmentations in ∼5 minutes. If the user has a FreeSurfer segmentation they would like to use, as specified in the previous section, the pipeline allows it as an input.

#### Electrode Region of Interest Assignment

Once an atlas has been chosen, the electrodes transformed into pre-implant MRI space can then be assigned to the specific ROIs in the pre-implant MRI. To do so, we generate a sphere with a user-defined radius around each electrode coordinate obtained from Module 2. The percent overlap of this sphere with the atlas brain regions is computed, and the region with the most overlap is the one assigned to that electrode. This process is repeated for each electrode. Besides identifying the region with the highest overlap, the software also generates a ranked list of percent overlaps when an electrode comes into contact with multiple regions. This feature enables users to choose a specific sphere-ROI overlap threshold that best aligns with their research question, offering more flexibility in the analysis process.

#### Template Space Registration

We provide the option for obtaining coordinates in MNI space after a rigid registration applied between the native space reference MRI and the MNI152NLin2009cAsym template. MNI registration is solely for visualization purposes and the outputs of this portion of the pipeline should not be used for electrode localization within ROIs.

### Post-Surgery and Post-Implant Registration

Our pipeline additionally provides the option for registering a post-surgery MRI (e.g. an MRI with a surgical resection cavity) to the original pre-implant MRI. This is useful to detect the electrodes that were implanted in resected brain areas. The rigid registration between the post-resection MRI and the pre-implant MRI uses the same Greedy parameters as the one used for registering the post-implant CT to pre-implant MRI. An example output from this post-resection registration pipeline is shown in **Figure 3**. Following similar steps, our pipeline can also register the post-implant MRI to the pre-implant MRI.

**Figure 3.**
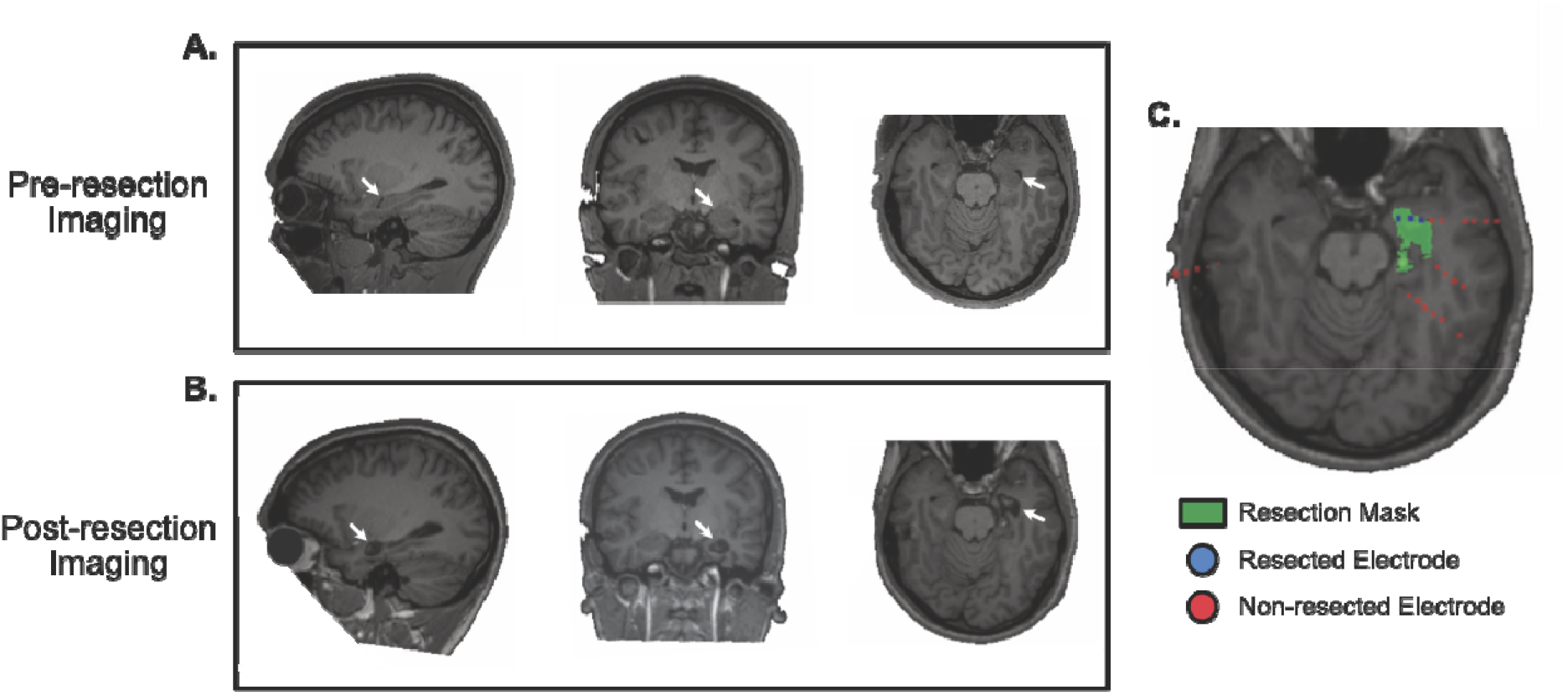
Estimating the original electrodes located in the post-surgical resection mask: Pre-resection MRI for an example subject who underwent resective surgery (**A.**), post-resection MRI (**B.**), and the Module 3 pipeline output identifying electrode contacts within the resected brain region (**C.**).

### Running iEEG-recon

We provide detailed documentation on how to run *iEEG-recon* in both Matlab and Python (https://ieeg-recon.readthedocs.io). To run the pipeline, the user will need the following minimum requirements: labeled electrode coordinates from VoxTool, a pre-implant MRI, and a post-implant CT. The process also utilizes a BIDS-like structure, which organizes the data for each subject according to subject IDs (e.g., sub-PENN01) and sessions (e.g., ses-research3T, ses-clinical01) using a BIDS-like naming convention. Semi-automatic electrode labeling with VoxTool takes between 20-35 minutes with a 10-16 electrode contact surgical plan.

For example, let us assume there is a subject called sub-PENN01, with pre-implant MRI stored in the session ses-research3T. The labeled electrode coordinates from VoxTool and the post-implant CT should be stored under the session ses-clinical01. This folder structure is illustrated in **Figure 7**.

Modules 2 and 3 from *ieeg_recon* with Greedy as a registration software and ANTsPyNet DKT segmentations can be executed in Python as follows:

`python ieeg_recon.py -d ∼/Desktop/BIDS/ -s sub-PENN01 -rs ses-research3T -cs ses-clinical01 -m -1 -gc -apn -mni -r 2`

- `d:` specifies the BIDS directory where all the subjects are located
- `s:` specifies the name of the subject
- `rs:` specifies the session name where the reference MRI is located
- `cs:` specifies the session name where the post-implant CT and the electrode coordinates from VoxTool are located
- `m:` specifies the module to run (-1 runs both modules 2 and 3)
- `gc:` specifies to run using Greedy
- `apn:` (optional) specifies to run ANTsPyNet DKT and Atropos segmentation for module 3
- `mni:` (optional) specifies to run an additional MNI registration in module 3 for visualization purposes
- `r:` specifies the radius (in mm) of the electrode spheres used to assign regions to each electrode coordinate

### Cloud Implementation

In order to scale clinical and research workflows, we leverage the capabilities of Pennsieve (https://app.pennsieve.io/), an open-source platform for data sharing, and Amazon Web Services (AWS) technologies. Pennsieve facilitates cross-collaborator data sharing by utilizing AWS’s S3 backend service. By deploying S3 as its backend storage, Pennsieve eliminates technological hurdles that collaborators may encounter when processing data on-site, thus enabling unrestricted access to data for processing.

To automate the iEEG-recon pipeline on a cloud infrastructure, we deploy our code on an AWS EC2 (Amazon Elastic Compute Cloud) instance. For our purposes, we regard an EC2 instance (https://docs.aws.amazon.com/ec2/) as a virtual machine capable of running the dockerized iEEG-recon software consistently across all collaborators. This approach simplifies the process and ensures uniformity, regardless of where the collaborator is based.

Our automation process involves the incorporation of AWS Lambda and Eventbridge technologies (https://docs.aws.amazon.com/lambda/). AWS Lambda, a powerful service for automating interactions between different AWS technologies, works in conjunction with Eventbridge, a user-friendly tool for constructing comprehensive pipelines using services like Lambda and EC2. This means that the long-term viability of the project is ensured even without a data engineer to create and maintain viable workflows.

We streamline the automation process as follows: First, data is uploaded to a public S3 bucket managed by the Pennsieve API. Eventbridge then monitors the S3 bucket for incoming data and applies a user-defined filter to preclude the initiation of an EC2 instance and prevent unnecessary costs. If Eventbridge identifies a match, it activates an EC2 instance. Within the EC2 environment, we compute the checksum of the new data and cross-check the output S3 bucket for any identical checksum values. If the data is unique, we run the docker container on the data, calculate the resulting data’s checksum, and push the data and its checksum to the output S3 bucket. The EC2 instance then closes. By running the pipeline only as needed, we can keep user cost to a minimum. Eventbridge is also capable of deciding when events run. This allows us to define set times when the pipeline runs or allows AWS to schedule the task as low priority for reduced cost.

The combination of an easy-to-use event scheduler, flexible pricing, and our dockerized container, we believe our approach allows for a scalable processing pipeline that is viable for institutions of all sizes. Final implementation of this cloud-based approach would require users to upload only anonymized data to avoid inadvertent sharing of private health information.

## Results

### Registration and Electrode ROI Assignment Within Minutes

One of the advantages of our pipeline is that the entirety of Modules 2 and 3, that is, the post-implant CT to pre-implant MRI registration and the electrode ROI assignment, run in about 10±4 minutes if Greedy is used for registration and ANTsPyNet is used for segmentation (default options). We tested the run time on standard laptops with a minimum of 8GB of RAM. This allows for fast turnaround times in situations where electrode reconstruction results are needed urgently (e.g. when a patient is being presented in a surgical conference), or when many subjects are being processed simultaneously from raw input images.

### Reconstructed Electrode Locations

Reconstructed electrode locations in MNI space (**Figure 4**) demonstrate a spatial bias for the temporal lobes, and specifically, the left temporal lobe. This is consistent with our studied population of mostly temporal lobe epilepsy patients. Medio-dorsal and medio-ventral structures were rarely implanted in our cohort.

**Figure 4.**
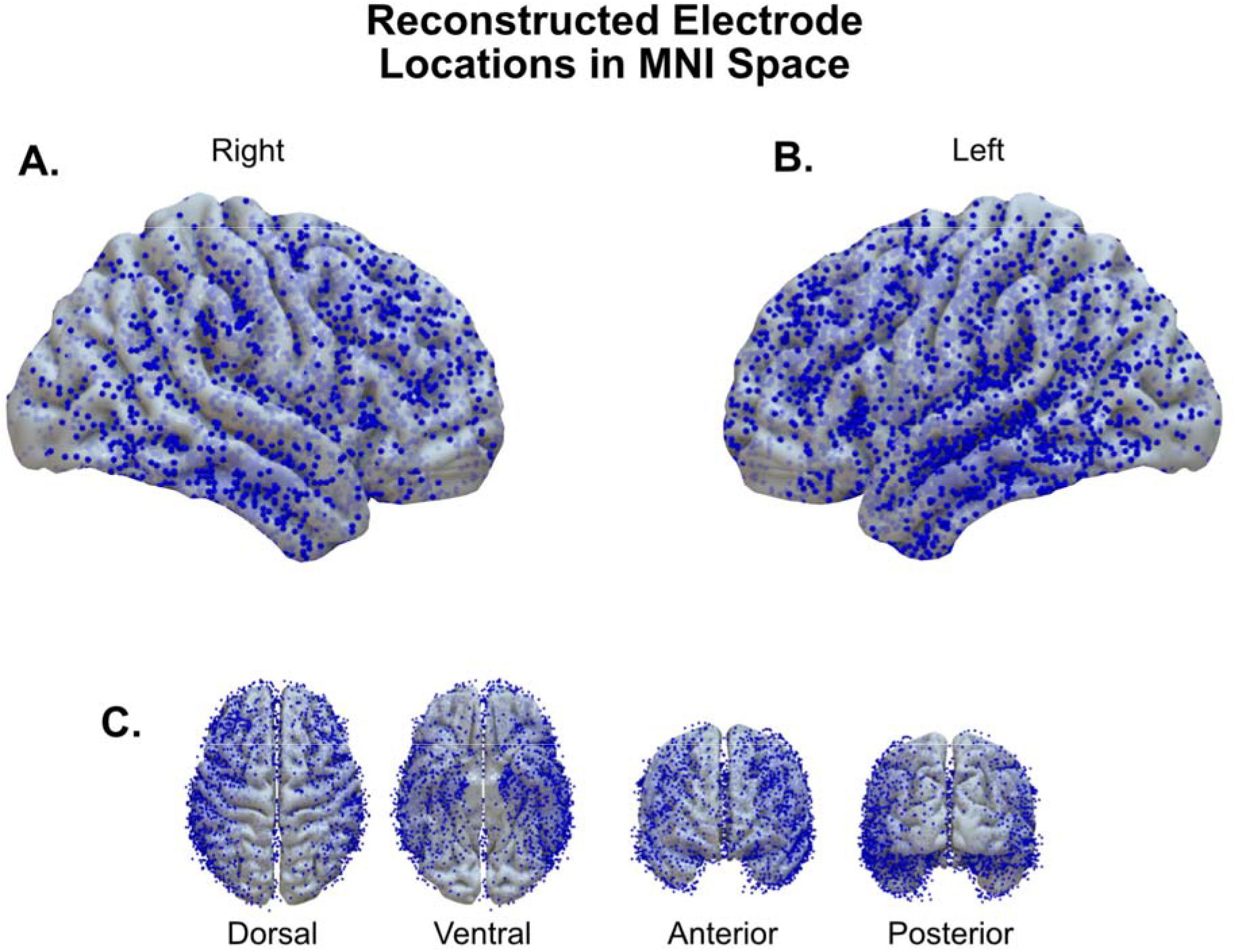
Reconstructed electrode locations in MNI template space: Electrode locations in MNI template space across all 98 subjects. Electrodes are represented as dots overlaid on an MNI surface template.

Across all patients, 9.2 ± 8.4% of the labeled electrodes were found to be in no tissue (e.g. outside the brain), 14.4 ± 8.5% in the CSF, 32.7 ± 11.7% in the gray matter, and 42.6 ± 13.5% in the white matter when using a 2mm radius for region assignment (**Figure 5A**). By default, *ieeg_recon* assigns regions based on a majority voting procedure, where the region with the most overlap with a 2mm sphere around the electrode radius is assigned to that electrode. However, sometimes brain shifts and post-surgical swelling can slightly alter the assignment of the electrodes. To overcome this, we provide a detailed JSON file that describes the percent overlap of each electrode sphere with all the regions it overlaps with, allowing for re-assignment of electrodes to different regions depending on different thresholds of overlap. In **Figure 5B-E** we show how the distribution of electrodes changes if we re-assign electrodes originally assigned to either white matter, CSF or outside the brain, to a gray matter region if their overlap with that region was at least as large as the specified threshold.

**Figure 5.**
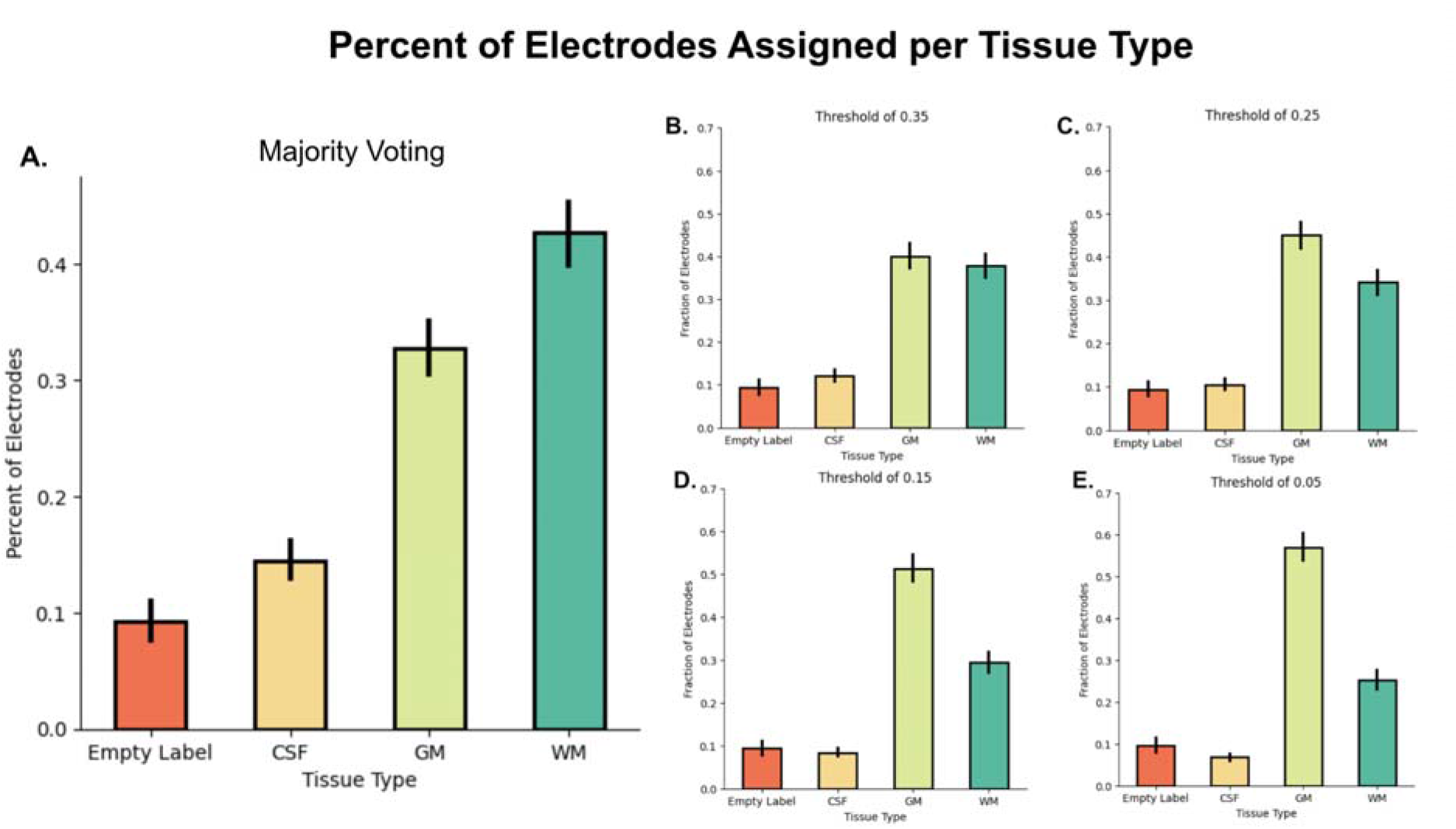
Percent of electrodes assigned to each tissue type: **Panel A.** Shows the percent of electrodes assigned to an empty label (outside of the brain), cerebrospinal fluid (CSF), gray matter (GM) and white matter (WM) across all subjects using a majority voting approach, that is, the region that had the largest overlap with the electrode sphere was assigned to that electrode. **Panels B-E** show the distribution of electrode assignments if the regions originally assigned to Empty Label, CSF or WM, was reassigned to a GM region if the GM overlap was above the specified threshold of 0.35 (**B.**), 0.25 (**C.**), 0.15 (**D.**), and 0.05 (**E.**).

Among the gray matter electrodes, we found that the top 3 implanted regions were the left middle, inferior and superior temporal gyri, with 71/98, 59/98 and 54/98 subjects having those regions implanted (**Table 2**). Similarly, when counting the number of gray matter electrodes implanted in each region, 373/4636 were found in the left middle temporal gyrus, 335/4636 in the left superior temporal gyrus, and 215/4636 in the left inferior temporal gyrus (**Table 3**).

**Table 2.**
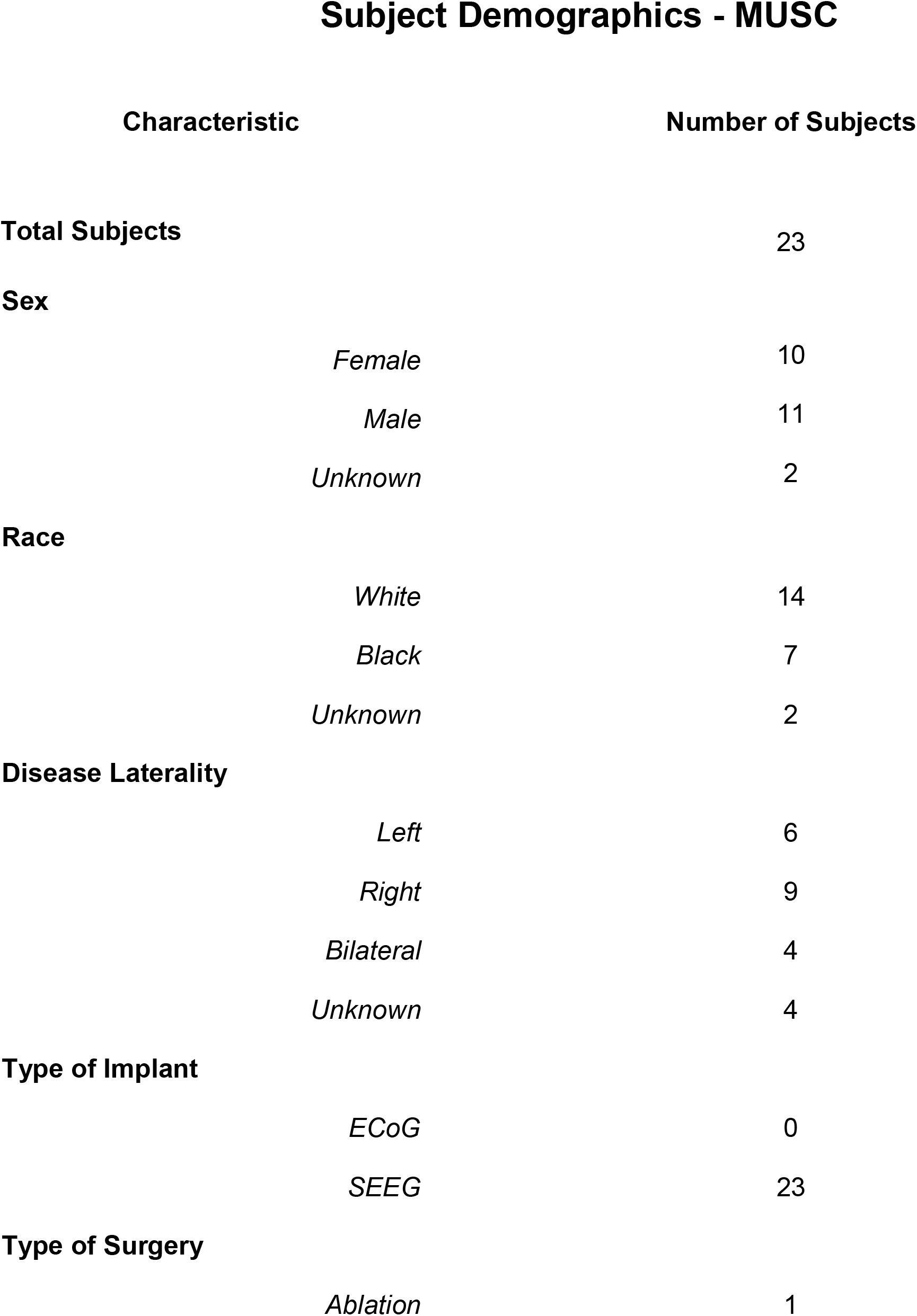
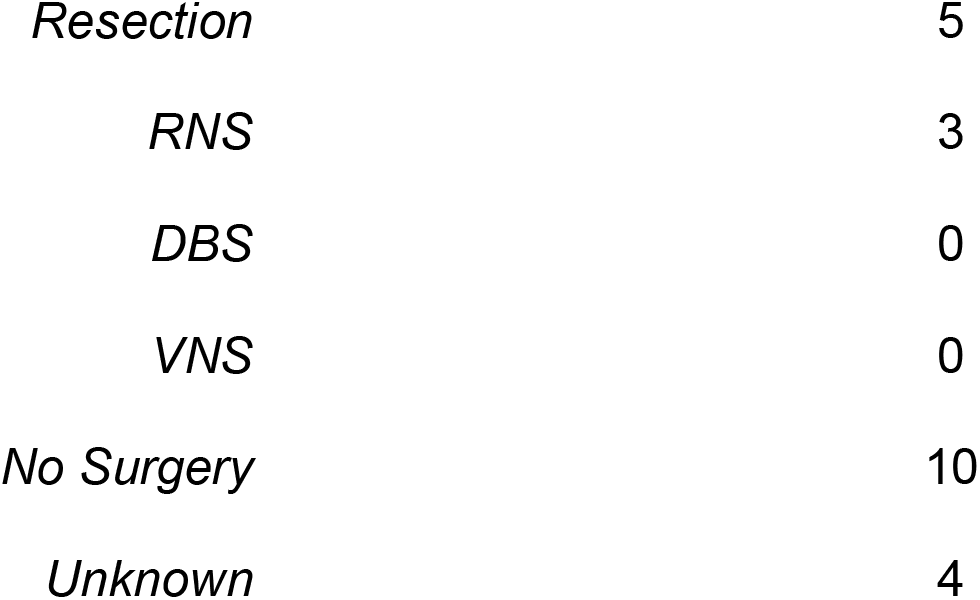
MUSC Subject demographics and clinical characteristics.

**Table 2a.**
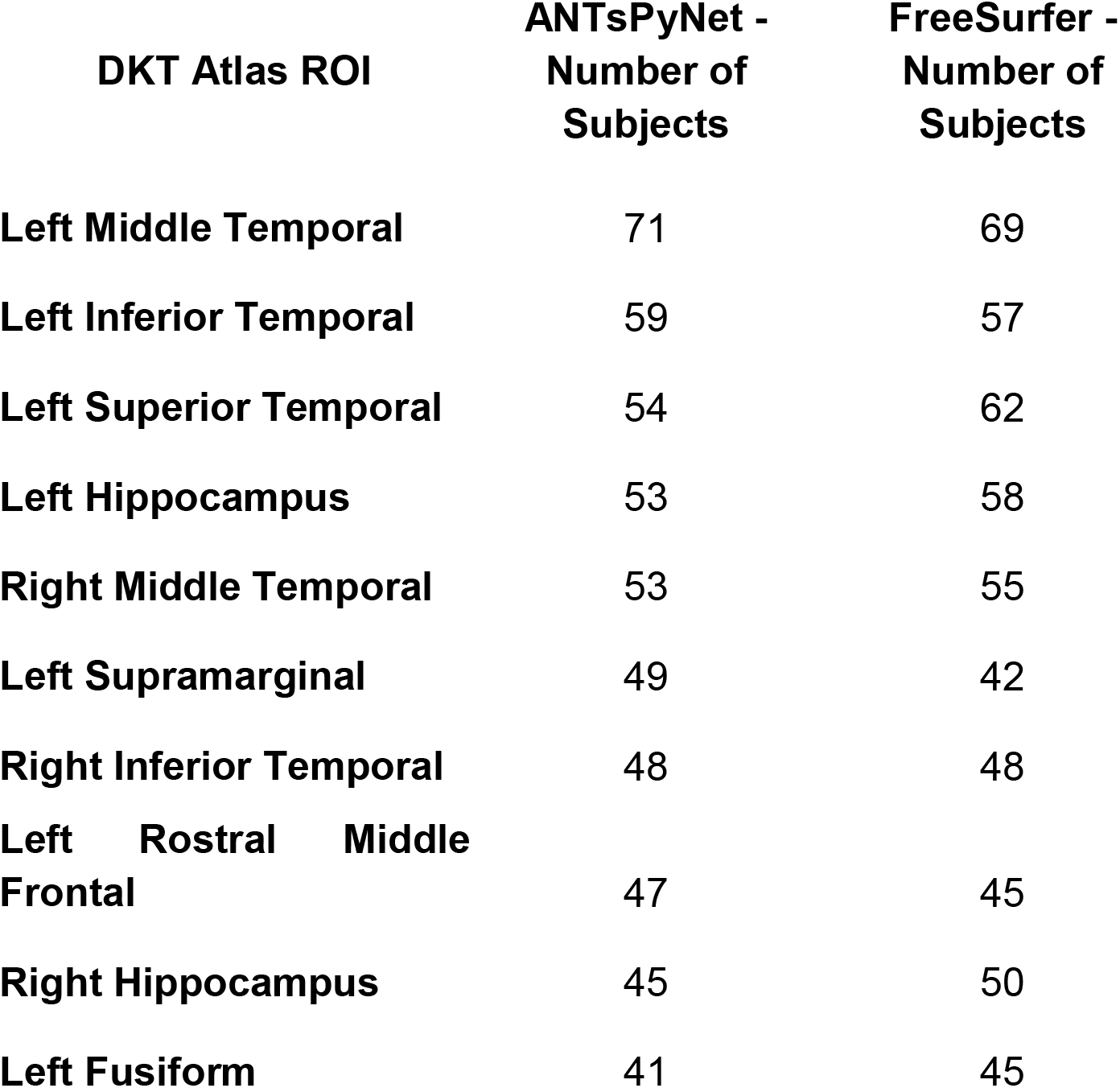
Number of subjects with at least one electrode per DKT ROIs.

**Table 3.**
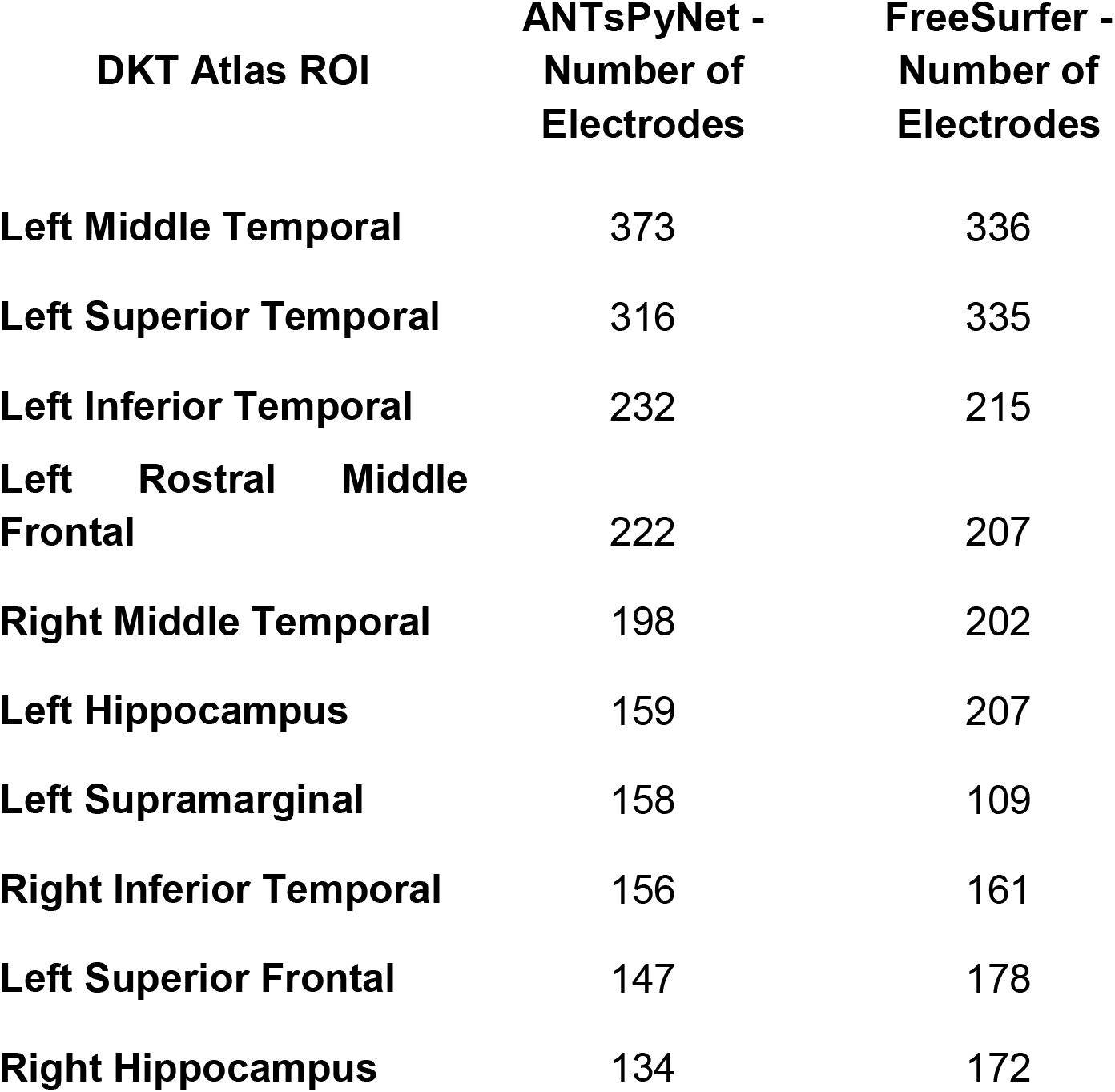
Number of electrodes assigned per DKT ROI.

| **DKT Atlas ROI** | **ANTsPyNet -**<br>**Number of**<br>**Electrodes** | **FreeSurfer -**<br>**Number of**<br>**Electrodes** |
| --- | --- | --- |
| **Left Middle Temporal** | 373 | 336 |
| **Left Superior Temporal** | 316 | 335 |
| **Left Inferior Temporal** | 232 | 215 |
| **Left Rostral Middle**<br>**Frontal** | 222 | 207 |
| **Right Middle Temporal** | 198 | 202 |
| **Left Hippocampus** | 159 | 207 |
| **Left Supramarginal** | 158 | 109 |
| **Right Inferior Temporal** | 156 | 161 |
| **Left Superior Frontal** | 147 | 178 |
| **Right Hippocampus** | 134 | 172 |

The electrode localizations for all patients were validated by visual inspection of post-implant MRI by a board-certified neuroradiologist (J.S.), which provides confidence in the accuracy and reliability of our pipeline.

### ANTsPyNet Electrode ROI Assignments are Comparable to FreeSurfer

The deep learning-based DKT segmentation provided by ANTsPyNet allows *ieeg_recon* to run quickly, while still providing accurate electrode ROI assignments. However, the most common approach for individualized brain segmentation in electrode reconstruction is to use FreeSurfer derived DKT segmentations^2–7^. We tested how consistent the electrode region assignments were between FreeSurfer and ANTsPyNet segmentations. At a group level, we found that the number of patients with electrodes implanted in each region was highly correlated between ANTsPyNet and FreeSurfer segmentations (Pearson’s *r=0.96*) (**Figure 6**). A similar correlation is seen if the number of electrodes assigned to each region across all patients is counted. However, the FreeSurfer implementation took an average of 5 hours on a dedicated computing server, whereas the ANTsPyNet approach was completed in an average of 10 minutes on the same server.

**Figure 6.**
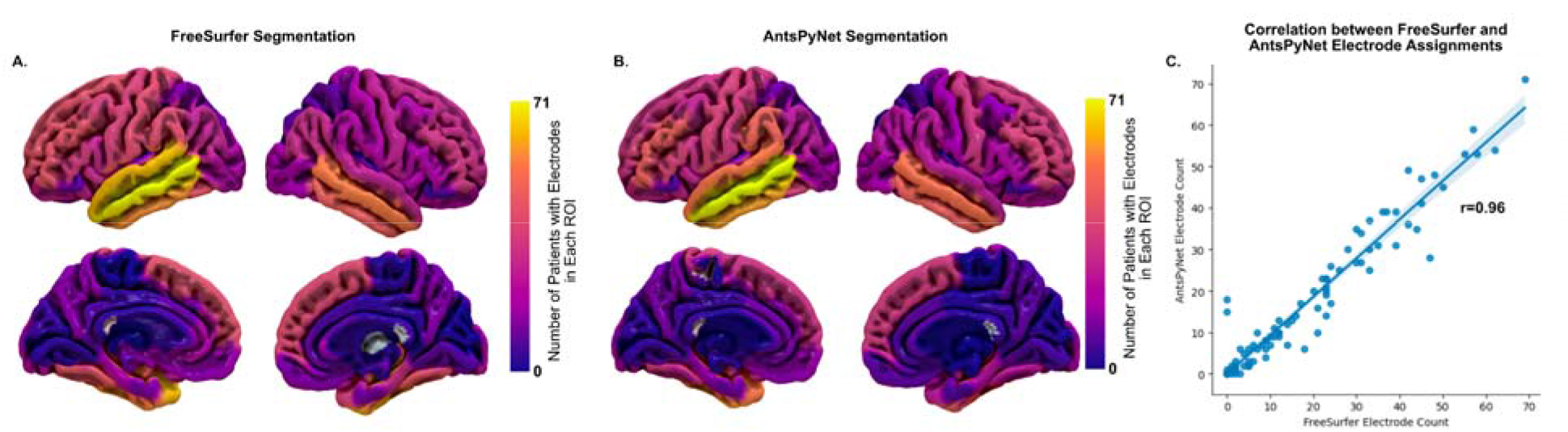
FreeSurfer and ANTsPyNet Desikan-Killany-Tourville atlas electrode assignments across subjects: **Panels A-B**. show the number of subjects for which an electrode was assigned to each of the regions in the Desikan-Killany-Tourville (DKT) atlas when the atlas segmentation was done by FreeSurfer (**A.**) or ANTsPyNet (**B.**). **Panel C** shows the correlation of that count across ROIs for the FreeSurfer and ANTsPyNet segmentations. *r – Pearson’s r*

**Figure 7.**
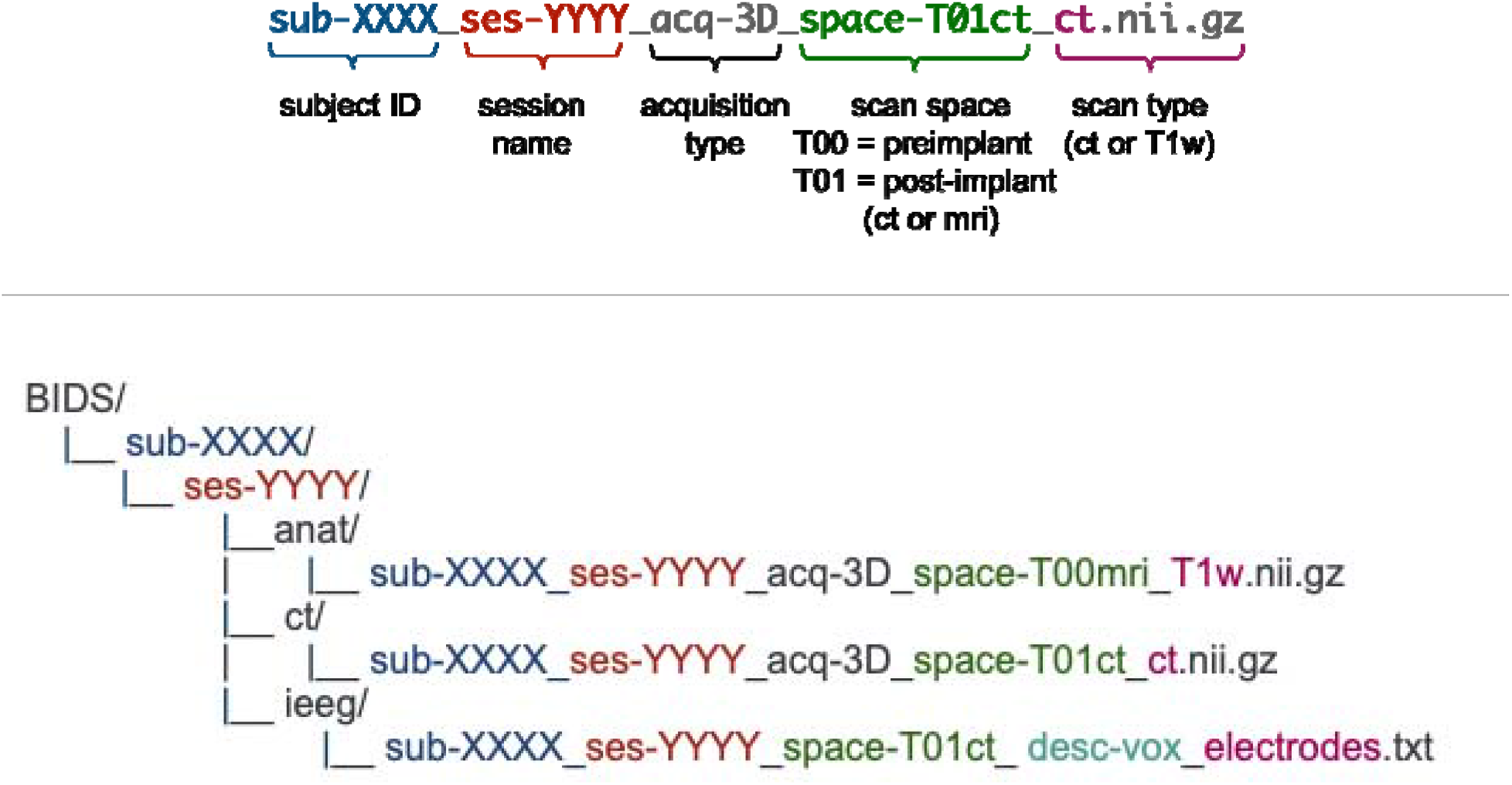
Folder Structure for iEEG-recon inputs.

### iEEG-recon is robust to implantable device artifact

Nine of the 98 patients were implanted with a responsive neurostimulation (RNS, NeuroPace Inc.) device subsequent to their iEEG implantation. The RNS computer is housed in a titanium case that is implanted within the thickness of the skull, and two electrode leads are implanted in the tissue of the brain. We used *ieeg-recon* to localize both the RNS electrode contacts and location of the device case by registering a post-RNS-implant CT scan to the pre-implant MRI. The post-RNS-implant CT scan presented a unique registration challenge due to the device artifact in the skull, since the registration steps operate by aligning the contours of the skull between CT and MRI images. For all RNS patients, we found that the pipeline outputs were acceptable as verified by visual inspection using our quality assessment reports and confirmed by a board-certified neuroradiologist (J.S.). The successful electrode localization in these RNS patients demonstrates the versatility of our reconstruction pipeline in handling multiple types of potential imaging artifacts.

## Discussion

We present iEEG-recon, an innovative electrode reconstruction pipeline that leverages cutting-edge image registration and segmentation algorithms for rapid and precise intracranial electrode localization. iEEG-recon’s scalable modular design enables users to select the most suitable options for their research objectives or clinical applications. We evaluated and tested iEEG-recon in a cohort of retrospective and prospective patients with drug-resistant epilepsy who underwent iEEG implantations at two epilepsy centers. The tool’s accuracy, speed, and compatibility with cloud platforms, demonstrated through its deployment on the Flywheel and Pennseive data infrastructure, make it a valuable resource for epilepsy centers worldwide. Furthermore, iEEG-recon proved robust against artifacts caused by RNS, highlighting its utility for patients with intracranial devices. In summary, iEEG-recon demonstrates the first step towards developing, validating, and testing scalable quantitative tools for standardized data analysis and seamless integration into clinical workflows, for advancing epilepsy treatment through multicenter collaborations.

Current electrode reconstruction pipelines^3, 4, 7^ have several limitations that hinder their widespread adoption across clinical sites that we believe iEEG-recon addresses. First, there is a steep learning curve for many of these tools, requiring the user to perform scripting tasks or to directly write code in a specific programming language in order to run the pipeline. iEEG-recon is self-contained, and as long as the input files are in the correct format and the electrodes have been labeled, the rest of the pipeline runs with a single command. Second, many of these tools are not scalable, as they require complex setups, and sometimes closed source software, limiting flexibility and modularity. Providing a Docker container allows iEEG-recon to be executed in any machine or server capable of running Docker, allowing the exact same software to be executed across multiple clinical or research sites. Finally, many currently available pipelines make use of effective, yet old, registration and segmentation technology, which leads to high computational requirements and running times. By leveraging advances in image registration and deep-learning based segmentation, iEEG-recon allows for complete electrode reconstruction and region-of-interest electrode assignments within 10-15 minutes, a significant improvement relative to current approaches.

Electrode reconstruction remains a critical component of presurgical epilepsy planning^22^, and iEEG-recon can help streamline and improve this process. iEEG-recon could be used to accurately localize intracranial electrodes in patients with refractory epilepsy, providing important information for surgical planning, such as where the seizure-onset zone (SOZ) electrodes are located^23^, and whether there is overlap with inoperable structures such as the eloquent cortex. We tested iEEG-recon with both ECOG and SEEG. While brain shifts—the inward displacement of brain tissue and electrodes due to pressure changes related to the craniotomy—were frequently observed with ECOG, they seldom occur in SEEG^24, 25^. To address these shifts, electrode positions may be projected to align with the brain surface’s normal vector. We do not use explicit surface maps to project ECOG grids onto the pre-implant cortical surface in iEEG-recon. Although this could be incorporated, it may not be necessary due to the growing prevalence of SEEG and lesser incidence of brain shifts on SEEG. Furthermore, our majority voting strategy already mitigates the impact of brain shift. After a patient has undergone epilepsy surgery, iEEG-recon could be used to confirm whether the post-surgical resection site overlaps with the suspected SOZ electrodes. iEEG-recon can also be used to identify the location of RNS electrodes for post-surgical confirmation.

Electrode reconstruction is also critical for research that involves intracranial electrophysiology^26^. Many intracranial electrophysiology studies focus on specific brain regions, such as the hippocampus^27, 28^ and the motor cortex^29, 30^, in order to understand function in health and disease. Therefore, accurate identification of electrodes within the target regions is necessary to generate reproducible findings, and to ensure that the measured signal is associated with the structure of interest. Multimodal studies that combine iEEG with neuroimaging, such as diffusion tensor imaging, are also of interest across disciplines, including epilepsy^12, 31, 32^. Functional neuroimaging, such as resting-state fMRI, has also demonstrated widespread abnormalities in focal epilepsy^33–35^, yet direct intracranial electrophysiological correlates of these abnormalities are lacking. iEEG-recon provides a natural framework for bridging intracranial electrophysiology and neuroimaging by allowing different structural and functional neuroimaging based atlases to be used in the electrode reconstruction process.

The use of federated data analysis has emerged as a promising approach in neuroimaging and iEEG studies, enabling researchers to perform large-scale multi-center analysis while maintaining data privacy^36–38^. We built iEEG-recon to work with standardized data formats (BIDS) and to streamline the processing of neuroimaging data, reducing the need for specialized technical expertise and enabling researchers to focus on interpreting their results. By improving the accessibility of neuroimage processing, our pipeline can facilitate the collaboration of multiple research teams across centers, ultimately leading to more robust and generalizable findings^39^.

There are several limitations of our current work. First, while iEEG-recon has been successfully tested in two level 4 adult epilepsy centers, its efficacy in pediatric populations remains unknown. Second, while self-contained, iEEG-recon leverages advances in image registration and segmentation algorithms, which have been sourced from other toolboxes that may still be under development. While these advancements have enabled the tool to provide more efficient results, it is important to consider the potential limitations of these technologies. Finally, it is important to mention that iEEG-recon is currently not FDA approved, and further testing and optimization are needed before it can be implemented reliably in clinical practice. However, the authors invite collaboration and feedback from the scientific and medical communities to improve and refine the tool’s performance.

## Conclusion

Reconstructing iEEG electrodes and precisely localizing them is necessary for research and clinical applications. A primary goal of our open-access tool is to make electrode reconstruction accessible to those with limited computing background or resources. Future work should iterate upon our foundation and we envisage improvement of our tool with widespread use and feedback.

## Data Availability

Our pipeline, with example data, is available here: https://ieeg-recon.readthedocs.io

https://github.com/penn-cnt/ieeg-recon

## Data Availability

Our pipeline, with example data, is available here: https://ieeg-recon.readthedocs.io

https://github.com/penn-cnt/ieeg-recon

## Funding

AL and KAD received support from NINDS (R01NS116504). NS received support from American Epilepsy Society (953257) and NINDS (R01NS116504). The authors would also like to thank the Thornton Foundation for their generous support. Brian Litt acknowledges funding from the Pennsylvania Tobacco Fund, NINDS R56099348, NIH DP1NS122038, the Mirowski Family Foundation, Jonathan Rothberg, and Neil and Barbara Smit.

## Notes

### Competing Interest Statement

The authors have declared no competing interest.

### Author Declarations

Patients were enrolled serially between 2015 and 2023 after providing written informed consent for iEEG data analysis, in line with the University of Pennsylvania's IRB-approved protocol (reference number 821778).

## References

1. World Health Organization. Epilepsy: A Public Health Imperative: Summary. World Health Organization; 2019:12 p.

2. Li G, Jiang S, Chen C, et al. iEEGview: an open-source multifunction GUI-based Matlab toolbox for localization and visualization of human intracranial electrodes. J Neural Eng. 2019;17(1):016016. doi:10.1088/1741-2552/ab51a5

3. Blenkmann AO, Phillips HN, Princich JP, et al. iElectrodes: A Comprehensive Open-Source Toolbox for Depth and Subdural Grid Electrode Localization. Front Neuroinformatics. 2017;11. Accessed April 10, 2023. https://www.frontiersin.org/articles/10.3389/fninf.2017.00014

4. Groppe DM, Bickel S, Dykstra AR, et al. iELVis: An open source MATLAB toolbox for localizing and visualizing human intracranial electrode data. J Neurosci Methods. 2017;281:40–48. doi:10.1016/j.jneumeth.2017.01.022

5. Rockhill AP, Larson E, Stedelin B, et al. Intracranial Electrode Location and Analysis in MNE-Python. J Open Source Softw. 2022;7(70):3897. doi:10.21105/joss.03897

6. Princich J, Wassermann D, Latini F, et al. Rapid and efficient localization of depth electrodes and cortical labeling using free and open source medical software in epilepsy surgery candidates. Front Neurosci. 2013;7. Accessed April 10, 2023. https://www.frontiersin.org/articles/10.3389/fnins.2013.00260

7. Hamilton LS, Chang DL, Lee MB, Chang EF. Semi-automated Anatomical Labeling and Inter-subject Warping of High-Density Intracranial Recording Electrodes in Electrocorticography. Front Neuroinformatics. 2017;11. Accessed April 10, 2023. https://www.frontiersin.org/articles/10.3389/fninf.2017.00062

8. Jenkinson M, Smith S. A global optimisation method for robust affine registration of brain images. Med Image Anal. 2001;5(2):143–156. doi:10.1016/s1361-8415(01)00036-6

9. Jenkinson M, Bannister P, Brady M, Smith S. Improved Optimization for the Robust and Accurate Linear Registration and Motion Correction of Brain Images. NeuroImage. 2002;17(2):825–841. doi:10.1006/nimg.2002.1132

10. Yushkevich PA, Pluta J, Wang H, Wisse LEM, Das S, Wolk D. IC-P-174: Fast Automatic Segmentation of Hippocampal Subfields and Medial Temporal Lobe Subregions In 3 Tesla and 7 Tesla T2-Weighted MRI. Alzheimers Dement. 2016;12(7S_Part_2):P126-P127. doi:10.1016/j.jalz.2016.06.205

11. Yushkevich PA, Piven J, Hazlett HC, et al. User-guided 3D active contour segmentation of anatomical structures: significantly improved efficiency and reliability. NeuroImage. 2006;31(3):1116–1128. doi:10.1016/j.neuroimage.2006.01.015

12. Bernabei JM, Sinha N, Arnold TC, et al. Normative intracranial EEG maps epileptogenic tissues in focal epilepsy. Brain. Published online May 31, 2022:13. doi:https://doi.org/10.1093/brain/awab480

13. Desikan RS, Ségonne F, Fischl B, et al. An automated labeling system for subdividing the human cerebral cortex on MRI scans into gyral based regions of interest. NeuroImage. 2006;31(3):968–980. doi:10.1016/j.neuroimage.2006.01.021

14. Klein A, Tourville J. 101 Labeled Brain Images and a Consistent Human Cortical Labeling Protocol. Front Neurosci. 2012;6. Accessed April 10, 2023. https://www.frontiersin.org/articles/10.3389/fnins.2012.00171

15. Lancaster JL, Woldorff MG, Parsons LM, et al. Automated Talairach Atlas labels for functional brain mapping. Hum Brain Mapp. 2000;10(3):120–131. doi:10.1002/1097-0193(200007)10:3<120::AID-HBM30>3.0.CO;2-8

16. Rolls ET, Huang CC, Lin CP, Feng J, Joliot M. Automated anatomical labelling atlas 3. NeuroImage. 2020;206:116189. doi:10.1016/j.neuroimage.2019.116189

17. Schaefer A, Kong R, Gordon EM, et al. Local-Global Parcellation of the Human Cerebral Cortex from Intrinsic Functional Connectivity MRI. Cereb Cortex N Y N 1991. 2018;28(9):3095–3114. doi:10.1093/cercor/bhx179

18. Yushkevich PA, Pluta JB, Wang H, et al. Automated volumetry and regional thickness analysis of hippocampal subfields and medial temporal cortical structures in mild cognitive impairment. Hum Brain Mapp. 2015;36(1):258–287. doi:10.1002/hbm.22627

19. Su JH, Thomas FT, Kasoff WS, et al. Thalamus Optimized Multi Atlas Segmentation (THOMAS): fast, fully automated segmentation of thalamic nuclei from structural MRI. NeuroImage. 2019;194:272–282. doi:10.1016/j.neuroimage.2019.03.021

20. Revell AY, Silva AB, Arnold TC, et al. A framework For brain atlases: Lessons from seizure dynamics. NeuroImage. 2022;254:118986. doi:10.1016/j.neuroimage.2022.118986

21. Tustison NJ, Cook PA, Holbrook AJ, et al. The ANTsX ecosystem for quantitative biological and medical imaging. Sci Rep. 2021;11(1):9068. doi:10.1038/s41598-021-87564-6

22. Sinha N. Localizing epileptogenic tissues in epilepsy: are we losing (the) focus? Brain. 2022;145(11):3735–3737. doi:10.1093/brain/awac373

23. Sinha N, Dauwels J, Kaiser M, et al. Predicting neurosurgical outcomes in focal epilepsy patients using computational modelling. Brain. 2017;140(2):319–332. doi:10.1093/brain/aww299

24. Sweet JA, Hdeib AM, Sloan A, Miller JP. Depths and grids in brain tumors: Implantation strategies, techniques, and complications. Epilepsia. 2013;54(s9):66–71. doi:10.1111/epi.12447

25. Stolk A, Griffin S, Van Der Meij R, et al. Integrated analysis of anatomical and electrophysiological human intracranial data. Nat Protoc. 2018;13(7):1699–1723. doi:10.1038/s41596-018-0009-6

26. Bernabei JM, Li A, Revell AY, et al. Quantitative approaches to guide epilepsy surgery from intracranial EEG. Brain. Published online January 10, 2023:awad007. doi:10.1093/brain/awad007

27. Piai V, Anderson KL, Lin JJ, et al. Direct brain recordings reveal hippocampal rhythm underpinnings of language processing. Proc Natl Acad Sci. 2016;113(40):11366–11371. doi:10.1073/pnas.1603312113

28. Rutishauser U, Reddy L, Mormann F, Sarnthein J. The Architecture of Human Memory: Insights from Human Single-Neuron Recordings. J Neurosci. 2021;41(5):883–890. doi:10.1523/JNEUROSCI.1648-20.2020

29. Chartier J, Anumanchipalli GK, Johnson K, Chang EF. Encoding of Articulatory Kinematic Trajectories in Human Speech Sensorimotor Cortex. Neuron. 2018;98(5):1042–1054.e4. doi:10.1016/j.neuron.2018.04.031

30. Martin S, Millán J del R, Knight RT, Pasley BN. The use of intracranial recordings to decode human language: challenges and opportunities. Brain Lang. 2019;193:73–83. doi:10.1016/j.bandl.2016.06.003

31. Shah P, Ashourvan A, Mikhail F, et al. Characterizing the role of the structural connectome in seizure dynamics. Brain. 2019;142(7):1955–1972. doi:10.1093/brain/awz125

32. Sinha N, Duncan JS, Diehl B, et al. Intracranial EEG structure-function coupling predicts surgical outcomes in focal epilepsy. Published online April 17, 2022. doi:10.48550/arXiv.2204.08086

33. Lucas A, Mouchtaris S, Cornblath EJ, et al. Subcortical Functional Connectivity Gradients in Temporal Lobe Epilepsy. NeuroImage Clin. Published online May 5, 2023:103418. doi:10.1016/j.nicl.2023.103418

34. Boerwinkle VL, Cediel EG, Mirea L, et al. NetworkLJtargeted approach and postoperative restingLJstate functional magnetic resonance imaging are associated with seizure outcome. Ann Neurol. 2019;86(3):344–356. doi:10.1002/ana.25547

35. Lucas A, Cornblath EJ, Sinha N, et al. Resting state functional connectivity demonstrates increased segregation in bilateral temporal lobe epilepsy. Epilepsia. 2023;64(5):1305–1317. doi:10.1111/epi.17565

36. Larivière S, Royer J, Rodríguez-Cruces R, et al. Structural network alterations in focal and generalized epilepsy assessed in a worldwide ENIGMA study follow axes of epilepsy risk gene expression. Nat Commun. 2022;13(1):4320. doi:10.1038/s41467-022-31730-5

37. Plis SM, Sarwate AD, Wood D, et al. COINSTAC: A Privacy Enabled Model and Prototype for Leveraging and Processing Decentralized Brain Imaging Data. Front Neurosci. 2016;10. doi:10.3389/fnins.2016.00365

38. Scheid BH, Bernabei JM, Khambhati AN, et al. Intracranial electroencephalographic biomarker predicts effective responsive neurostimulation for epilepsy prior to treatment. Epilepsia. 2022;63(3):652–662. doi:10.1111/epi.17163

39. Sinha N, Johnson GW, Davis KA, Englot DJ. Integrating Network Neuroscience Into Epilepsy Care: Progress, Barriers, and Next Steps. Epilepsy Curr. 2022;22(5):272–278. doi:10.1177/15357597221101271

